# SARS-CoV-2 spike protein S1 induces fibrin(ogen) resistant to fibrinolysis: Implications for microclot formation in COVID-19

**DOI:** 10.1101/2021.03.05.21252960

**Authors:** Lize M. Grobbelaar, Chantelle Venter, Mare Vlok, Malebogo Ngoepe, Gert Jacobus Laubscher, Petrus Johannes Lourens, Janami Steenkamp, Douglas B. Kell, Etheresia Pretorius

## Abstract

Severe acute respiratory syndrome coronavirus 2 (SARS-Cov-2)-induced infection, the cause of coronavirus disease 2019 (COVID-19), is characterized by unprecedented clinical pathologies. One of the most important pathologies, is hypercoagulation and microclots in the lungs of patients. Here we study the effect of isolated SARS-CoV-2 spike protein S1 subunit as potential inflammagen *sui generis*. Using scanning electron and fluorescence microscopy as well as mass spectrometry, we investigate the potential of this inflammagen to interact with platelets and fibrin(ogen) directly to cause blood hypercoagulation. Using platelet poor plasma (PPP), we show that spike protein may interfere with blood flow. Mass spectrometry also showed that when spike protein S1 is added to healthy PPP, it results in structural changes to β and γ fibrin(ogen), complement 3, and prothrombin. These proteins were substantially resistant to trypsinization, in the presence of spike protein S1. Here we suggest that, in part, the presence of spike protein in circulation may contribute to the hypercoagulation in COVID-19 positive patients and may cause substantial impairment of fibrinolysis. Such lytic impairment may result in the persistent large microclots we have noted here and previously in plasma samples of COVID-19 patients. This observation may have important clinical relevance in the treatment of hypercoagulability in COVID-19 patients.

## INTRODUCTION

Severe acute respiratory syndrome coronavirus 2 (SARS-Cov-2)-induced infection, the cause of coronavirus disease 2019 (COVID-19), is characterized by unprecedented clinical pathologies. Phenotypic vascular characteristics are strongly associated with various coagulopathies that may result in either bleeding and thrombocytopenia or hypercoagulation and thrombosis (Gupta et al., 2020, Perico et al., 2021). Various circulating and dysregulated inflammatory coagulation biomarkers, including fibrin(ogen), D-dimer, P-selectin and von Willebrand Factor (VWF), C-reactive protein (CRP), and various cytokines, directly bind to endothelial receptors. Endotheliopathies are therefore a key clinical feature of the condition (Goshua et al., 2020, Ackermann et al., 2020). During the progression of the various stages of the COVID-19, markers of viral replication, as well as VWF and fibrinogen depletion with increased D-dimer levels and dysregulated P-selectin levels, followed by a cytokine storm, are likely to be indicative of a poor prognosis (Grobler et al., 2020, Pretorius et al., 2020, Venter et al., 2020, Roberts et al., 2020). This poor prognosis is further worsened as together with a substantial deposition of microclots in the lungs (Renzi et al., 2020, Ciceri et al., 2020, Bobrova et al., 2020), plasma of COVID-19 patients also carries a massive load of preformed amyloid clots (Grobler et al., 2020), and there are also numerous reports of damage to erythrocytes (Lam et al., 2020, Berzuini et al., 2020, Akhter et al., 2020), platelets and dysregulation of inflammatory biomarkers (Grobler et al., 2020, Pretorius et al., 2020, Venter et al., 2020, Roberts et al., 2020).

The virulence of the pathogen is closely linked to its membrane proteins. One such protein, found on the COVID-19 virus, is the spike protein, which is a membrane glycoprotein. The spike proteins are the key factors for virus attachment to target cells, as they bind to the angiotensin-converting 2 (ACE2) surface receptors (Bergmann and Silverman, 2020). Spike proteins are class I viral fusion proteins (Kawase et al., 2019). They present as protruding homotrimers on the viral surface and mediate virus entry into the target host cells (Walls et al., 2020). A singular spike protein is between 180–200 kDa in size and contains an extracellular N-terminal, a transmembrane domain fixed in the membrane of the virus, and a short intracellular C-terminal segment (Kawase et al., 2019, Zhang et al., 2020). Spike proteins are coated with polysaccharide molecules that serve as camouflage. This helps evade surveillance by the host immune system during entry (Zhang et al., 2020). The S1 subunit is responsible for receptor binding (Watanabe et al., 2020), with subunit 2 (S2), a carboxyl-terminal subunit, responsible for viral fusion and entry (Flores-Alanis et al., 2020) (see Figure 1).

**Figure 1:**
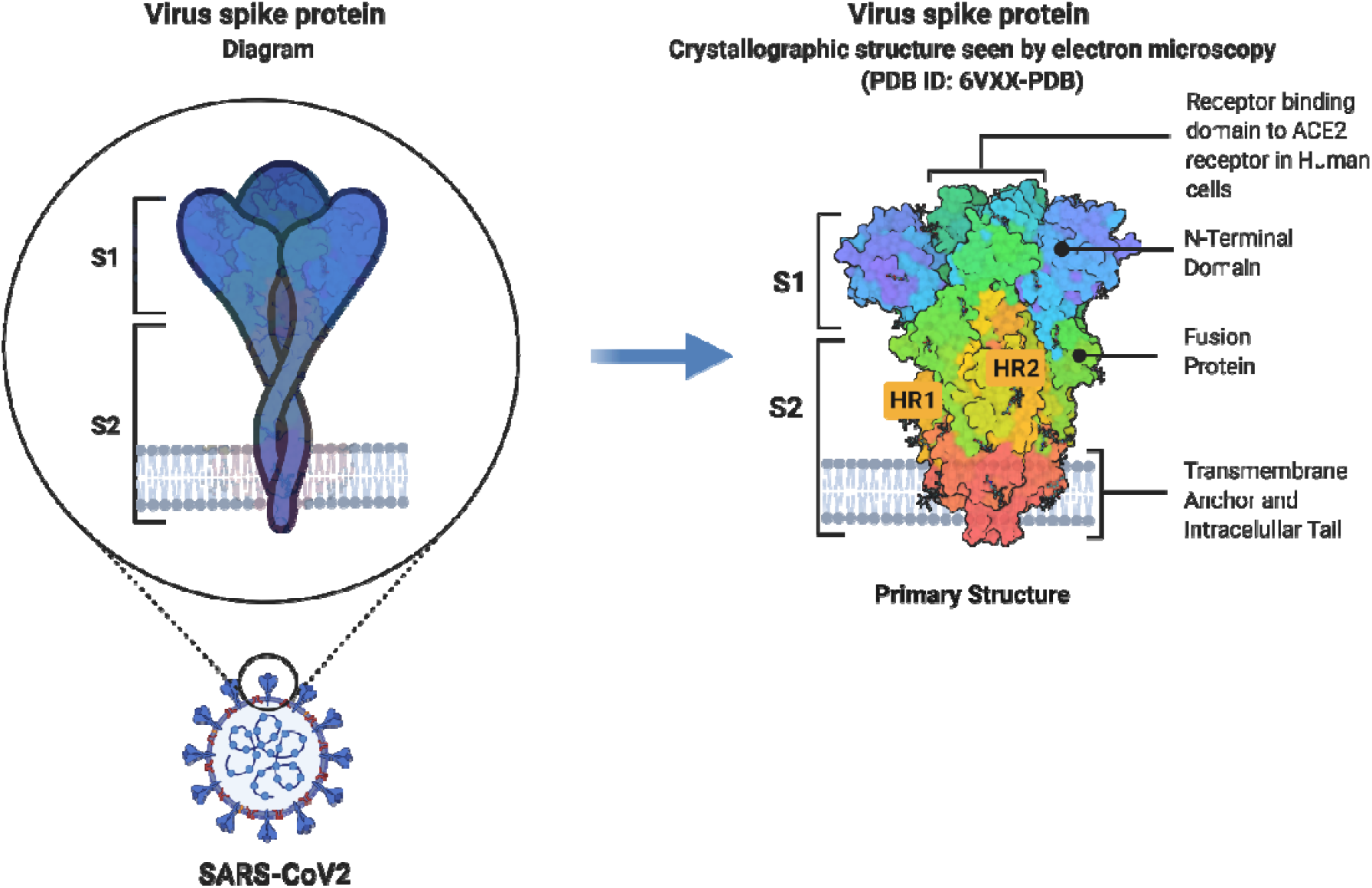
Schematic representation of SARS-CoV-2 Spike glycoprotein. Adapted from (Duan et al., 2020). **Abbreviations:** S1, subunit 1; S2, subunit 2; HR1, heptad repeat 1; HR2, heptad repeat 2. This image was created with BioRender (https://biorender.com/).

Receptor binding is certainly responsible for cell-mediated pathologies, but does not of itself explain the coagulopathies. Spike protein, can however be shed, and it has been detected in various organs, including the urinary tract (George et al., 2021). S1 proteins can also cross the blood-brain-barrier (Rhea et al., 2021). Free S1 particles may also play a role in the pathogenesis of the disease (Letarov et al., 2020, Buzhdygan et al., 2020). Free spike protein can potentially be released due to spontaneous “firing” of the S protein trimers on the surface of virions, and infected cells liberates free receptor binding domain-containing S1 particles (Letarov et al., 2020).

Here we study the effect of isolated SARS-CoV-2 spike protein S1 subunit as potential pro-inflammatory inflammagen *sui generis*. We investigate the potential of this inflammagen to directly interact with platelets and fibrin(ogen) to cause fibrin(ogen) protein changes and blood hypercoagulation. We also determine if the spike protein may interfere with blood flow, by comparing naïve healthy PPP samples, with and without added spike protein, to PPP samples from COVID-19 positive patients (before treatment). We conclude that the spike protein may have pathological effects directly, without being taken up by cells. This provides further evidence that targeting it directly, whether via vaccines or antibodies, is likely to be of therapeutic benefit.

## MATERIALS AND METHODS

### Ethical clearance

Ethical clearance for the study was obtained from the Health Research Ethics Committee (HREC) of Stellenbosch University (South Africa) (reference: N19/03/043, project ID: 9521). The experimental objectives, risks, and details were explained to volunteers both verbally and in text and informed consent were obtained prior to blood collection. Strict compliance to ethical guidelines and principles Declaration of Helsinki, South African Guidelines for Good Clinical Practice, and Medical Research Council Ethical Guidelines for Research were kept for the duration of the study and for all research protocols.

### Sample demographics and considerations

Blood was collected from healthy volunteers (N=11; 3 males, 8 females; mean age 43.4 ± 11.7) to serve as controls. Individuals who smoke, who were diagnosed with cardiovascular diseases, clotting disorders (coagulopathies), and/or any known inflammatory conditions (e.g. T2DM, rheumatoid arthritis, tuberculosis, asthma, etc.) could not serve as control volunteers. Furthermore, pregnancy, lactation, hormonal therapy, oral contraceptive usage, and/or using anticoagulants, were also factors that would result in exclusion. Smoking was excluded since it has been proven to impair coagulation, fibrinolysis, and the haemostatic process (Pretorius et al., 2010). Microfluidic analysis included a preliminary analysis using PPP samples from three COVID-19 positive patients, on day of first diagnosis and before any treatment were resumed. All three patients were diagnosed with moderate to severe COVID-19 symptoms (1 male; 2 females mean age 71 ± 14.1).

### Blood sample collection

Either a qualified phlebotomist or medical practitioner drew the volunteers’ (control) blood via venepuncture, adhering to standard sterile protocol. Blood samples were stored in two to three 4.5mL sodium citrate (3.2%) tubes (BD Vacutainer^®^, 369714). After several gentle inversions, the collected citrate tubes were allowed to rest at room temperature for a minimum of 30 minutes to allow for adequate anticoagulant amalgamation before commencing sample preparation. Whole blood (WB) was centrifuged at 3000xg for 15 minutes at room temperature to isolate erythrocytes. The supernatant, i.e. PPP, was collected and stored in 1.5mL Eppendorf tubes at −80°C, till needed for experiments.

### Spike protein preparation

SARS-CoV-2 (2019-nCoV) Spike protein S1 Subunit, mFcTag, was purchased from Sino Biological (Beijing, China) (catalog number 40591-V05H1) and prepared using doubly distilled water, following the instructions provided. 400µL diluent (endotoxin-free water) was added to the 100µg spike protein to create a stock solution (A) of 0.25mg.mL^-1^. This stock solution was diluted to working solutions. To determine the concentration of spike protein needed to result in significant, but yet a high enough concentration to cause physiological effects on the viscoelastic properties of blood, different concentrations of spike protein in PPP were assessed with fluorescence microscopy. A healthy control blood sample were separated into four 1.5mL Eppendorf tubes with different final exposure concentrations of spike protein in the PPP of 100ng.mL^-1^, 50ng.mL^-1^, 10ng.mL^-1^ and 1ng.mL^-1^. PPP samples were incubated with the various spike protein concentrations for 30 minutes at room temperature.

### Fluorescence microscopy of purified fibrinogen and platelet poor plasma (PPP) with and without thrombin

#### Concentration verification

To verify which spike protein concentration will be effective, 5uL of the PPP exposed to the varies spike protein concentrations was placed on a glass slide, after being exposure to the fluorescent amyloid dye, Thioflavin T (ThT) (Sigma-Aldrich, St. Louis, MO, USA) for 30 minutes at room temperature. The final concentration of ThT in all prepared samples was 0,005mM. After the evaluation of the samples, with the varies spike protein concentrations, it was found that the final exposure concentrations of 1ng.mL^-1^ was sufficient and used for the rest of the study.

#### Amyloid protein and anomalous clotting in platelet poor plasma samples

To study spontaneous anomalous clotting of fibrin(ogen), in the naïve healthy PPP samples, and in the presence of spike protein, 5 ⍰L PPP exposed to 1ng.mL^-1^ (final concentrations) of spike protein, was smeared on a glass slide and covered with a coverslip. This was done after it was exposure to the fluorescent amyloid dye, Thioflavin T (ThT) (final concentration: 0,005mM) (Sigma-Aldrich, St. Louis, MO, USA) for 30 minutes at room temperature. Fibrin PPP clots, with and without spike protein and after exposure to ThT, were also prepared by adding 2,5 ⍰L of thrombin (7U·mL^-1^, South African National Blood Service) to 5uL PPP and was placed on a glass slide and covered with a coverslip. The excitation wavelength for ThT was set at 450nm to 488nm and the emission at 499nm to 529nm and processed samples were viewed using a Zeiss Axio Observer 7 fluorescent microscope with a Plan-Apochromat 63x/1.4 Oil DIC M27 objective (Carl Zeiss Microscopy, Munich, Germany).

Fluorescence microscopy images of healthy PPP with and without spike protein were analysed using Fiji^®^ (Java 1.6_0 24 [64-bit]) to numerically represent the images. The total area of fluorescing particles or anomalous clotting (identified by the amyloid dye, ThT) (Kell and Pretorius, 2017, Pretorius et al., 2016, Pretorius et al., 2013b) was determined using a thresholding algorithm in Fiji^®^. Images were firstly set to scale in Fiji^®^ according to the magnification of the lens used on the fluorescent microscope, followed by the selection of appropriate threshold value to regard as much of the foreground and disregard as much of the background of the image as possible. In order to optimise the amount of images thresholded, a program was written in Java to simultaneously analyse a group of images (see supplementary material). The total percentage of anomalous clots in each image was calculated and the average of all the images per sample was calculated. These average values were used for statistical analysis.

#### Purified fibrin(ogen) clot model

To determine if spike protein causes changes in purified fibrinogen, our purified fibrin(ogen) clot model of choice was fluorescent fibrinogen conjugated to Alexa Fluor™488 (ThermoFisher, F13191). A final fibrinogen concentration of 2mg.mL^-1^ was prepared in endotoxin-free water and exposed to 1ng.mL^-1^ (final concentrations) spike protein for 30 minutes at room temperature. 5uL of the purified fibrinogen was placed on a glass slide, with 2,5uL of thrombin. The excitation wavelength for our fluorescent fibrinogen model was set at 450nm to 488nm and the emission at 499nm to 529nm and processed samples were viewed using a Zeiss Axio Observer 7 fluorescent microscope with a Plan-Apochromat 63x/1.4 Oil DIC M27 objective (Carl Zeiss Microscopy, Munich, Germany).

#### Whole blood

Healthy WB was exposed to a final exposure concentration of 1ng.mL^-1^ spike protein. The fluorescent marker, CD62P (platelet surface P-selectin) was added to WB to study platelet activation. CD62P is found on the membrane of platelets and then translocate to the platelet membrane surface. The translocation occurs after the platelet P-selectin is released from the cellular granules during platelet activation (Venter et al., 2020, Grobler et al., 2020). 4 µL CD62P (PE-conjugated) (IM1759U, Beckman Coulter, Brea, CA, USA) was added to 20 µL WB (either naïve or incubated with spike protein). The WB exposed to the marker was incubated for 30 minutes (protected from light) at room temperature. The excitation wavelength for the CD62P was 540nm to 570nm and the emission 577nm to 607nm. Processed samples were also viewed using a Zeiss Axio Observer 7 fluorescent microscope with a Plan-Apochromat 63x/1.4 Oil DIC M27 objective (Carl Zeiss Microscopy, Munich, Germany).

### Scanning electron microscopy of whole blood samples

Scanning electron microscopy (SEM) was used to view healthy WB samples, with and without the addition of spike protein. 10uL WB was placed on a glass cover slip and prepared according to previously published SEM preparation methods (Pretorius, 2013, Pretorius et al., 2013a), starting with washing steps in phosphate-buffered saline (PBS) (pH = 7.4) (ThermoFisher Scientific, 11594516) for 20 minutes. Fixation was performed by coating the slides in 4% formaldehyde (FA) for 30 minutes, followed by washing them in PBS three times. For each wash, the PBS should be left on for 3 minutes before removing and washing again. Osmium tetroxide (OsO_4_) (Sigma-Aldrich, 75632) was added for 15 minutes and the slides were washed in PBS three times with 3 minutes in each once more. The next step was to serially dehydrate the slides with ethanol followed by a drying step using hexamethyldisilazane (HMDS) (Sigma-Aldrich, 379212). Samples were mounted on glass slides and coated with carbon. The slides were viewed on a Zeiss MERLIN FE-SEM with the InLens detector at 1kV (Carl Zeiss Microscopy, Munich, Germany).

### Microfluidics

Microfluidic analysis was performed using healthy PPP and healthy pooled PPP samples (3 pooled PPP samples), with and without spike protein, and 3 COVID-19 PPP samples. Pooled samples were used due to the volume required for this experiment.

#### Hardware

A microfluidic setup was used to simulate and investigate clot growth under conditions of flow. A Cellix microfluidic syringe pump (Cellix Ltd, Dublin, Ireland) was used to drive flow through Cellix Vena8 Fluoro+ biochips (Cellix Ltd, Dublin, Ireland), comprising eight straight microfluidic flow channels each, at flow rates specified in the following paragraph. A single microchannel has a width of 400μm and a height of 100μm (equivalent diameter of 207μm), and a length of 2.8cm. The dimensions of the microchannel are in the same order of magnitude as those of some vessels of the microvasculature, that is, under 300μm (Jacob et al., 2016). In order to observe clot evolution in real-time, the biochips were placed under the Zeiss Axio Observer 7 fluorescent microscope with a 10x/0.25 objective (Carl Zeiss Microscopy, Munich, Germany).

#### Flow conditions

A new flow channel was used for every run. The channel was flushed with distilled water at a flow rate of 1mL.min^-1^ for 1 minute. After 5 minutes, thrombin was infused through the microchannel at a flow rate of 50 μL.min^-1^ for 90 seconds (Figure 2). The channel was left to stand for another five minutes and then the sample (control, control with spike, or COVID-19) was infused at a flow rate of 10μL.min^-1^ for 5 minutes, with a video recording of the channel (Figure 2). The flow was then stopped after 5 minutes, and a set of micrographs was taken across the channel. The sample was then left for another 5 minutes, to see if any additional changes occur (Figure 2). This flow rate corresponds with a shear rate, γ ≈ 250s^-1^ and a Reynolds numbers, Re ≤ 1. One of the main challenges in attempting to achieve consistent shear rates and Reynolds numbers was the variability in viscosity from sample to sample. Furthermore, blood flowing through microvessels is known to behave in a non-Newtonian manner, adding to the complexities of variable viscosity within a single sample. To achieve standardisation between samples, a constant flow rate was used.

**Figure 2:**
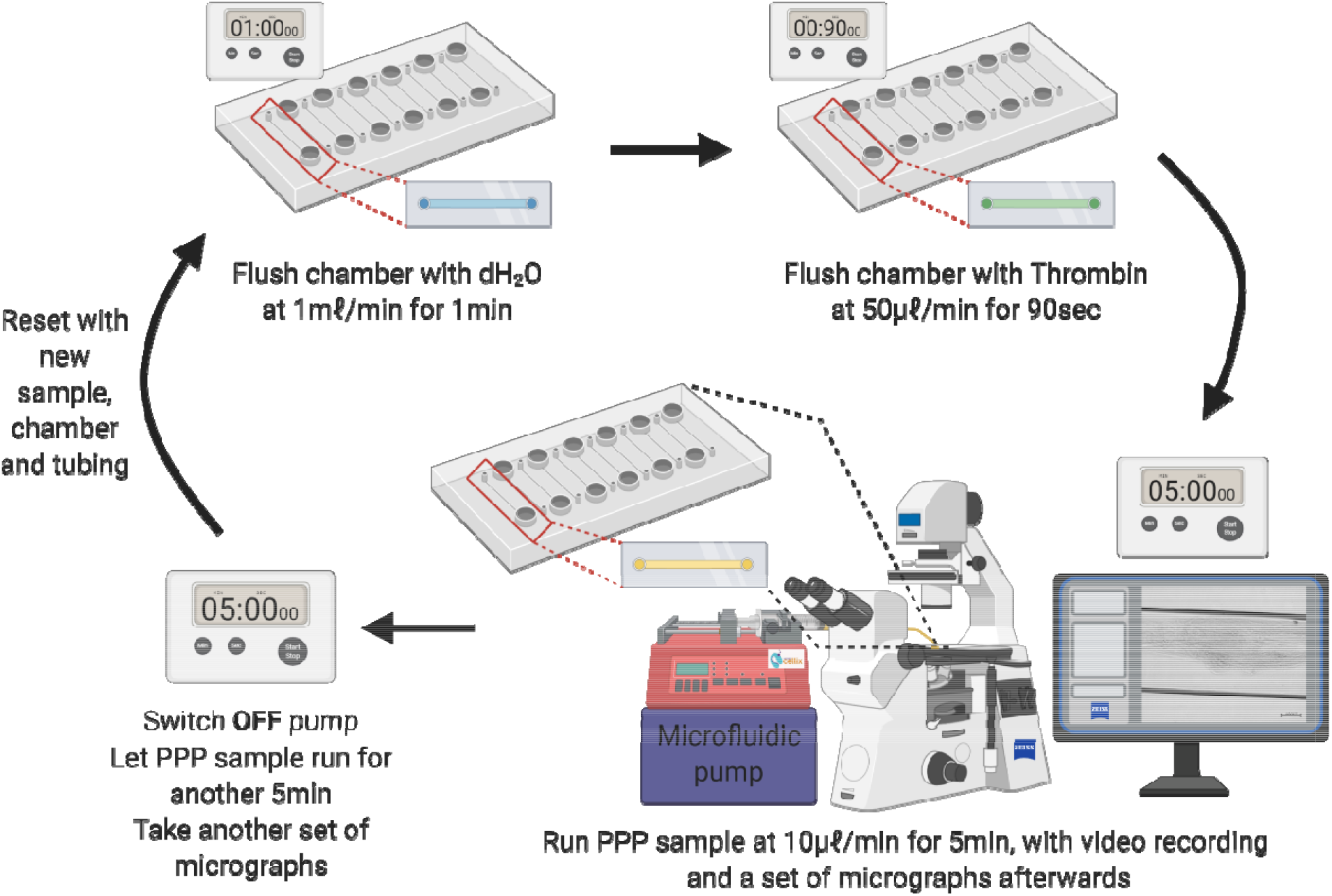
Experimental protocol for growing clots in a microfluidic system.

### Proteomics

Four healthy PPP samples were analysed before and after addition of spike protein. The samples were diluted in 10mM ammonium bicarbonate to 1mg.mL^-1^. A total of 1 ⍰g trypsin (New England Biosystems) was added to the plasma for 1:50 enzyme to substrate ratio. No reduction or alkylation was performed.

#### Liquid chromatography

##### Dionex nano-RSLC

Liquid chromatography was performed on a Thermo Scientific Ultimate 3000 RSLC equipped with a 5 mm × 300 µm C_18_ trap column (Thermo Scientific) and a CSH 25cmx75 µm 1.7 ⍰m particle size C_18_ column (Waters) analytical column. The solvent system employed was loading: 2% acetonitrile:water; 0.1% FA; Solvent A: 2% acetonitrile: water; 0.1% FA and Solvent B: 100% acetonitrile:water. The samples were loaded onto the trap column using loading solvent at a flow rate of 2 µL/min from a temperature controlled autosampler set at 7□C. Loading was performed for 5 minutes before the sample was eluted onto the analytical column. Flow rate was set to 300nL/minute and the gradient generated as follows: 5.0%-30% B over 60 minutes and 30-50%B from 60-80 minutes. Chromatography was performed at 45°C and the outflow delivered to the mass spectrometer.

##### Mass spectrometry

Mass spectrometry was performed using a Thermo Scientific Fusion mass spectrometer equipped with a Nanospray Flex ionization source. Plasma samples, before and after addition of spike protein addition (1 ng.mL^-1^ final exposure concentration), from 4 of our control samples were analysed with this method. The sample was introduced through a stainless-steel nano-bore emitter Data was collected in positive mode with spray voltage set to 1.8kV and ion transfer capillary set to 275°C. Spectra were internally calibrated using polysiloxane ions at m/z = 445.12003. MS1 scans were performed using the orbitrap detector set at 120 000 resolution over the scan range 375-1500 with AGC target at 4 E5 and maximum injection time of 50ms. Data was acquired in profile mode. MS2 acquisitions were performed using monoisotopic precursor selection for ion with charges +2-+7 with error tolerance set to +/-10ppm. Precursor ions were excluded from fragmentation once for a period of 60s. Precursor ions were selected for fragmentation in HCD mode using the quadrupole mass analyser with HCD energy set to 30%. Fragment ions were detected in the Orbitrap mass analyzer set to 30 000 resolution. The AGC target was set to 5E4 and the maximum injection time to 100ms. The data was acquired in centroid mode.

### Statistical analysis

#### Data analysis: plasma samples

Statistical analyses of data generated were performed using GraphPad Prism software (version 9.0.0). The normality of the data was assessed using the Shapiro-Wilk normality test. For analysis of data between two groups, paired *t*-tests (for pairwise statistical comparisons between data from untreated and treated control groups) and unpaired *t*-tests (for non-pairwise statistical comparisons) were performed to assess statistical significance for parametric data, whereas the Mann-Whitney test was utilized to test for statistical significance in non-parametric data and the Wilcoxon test was used for significance in paired parametric data. When comparing three or more experimental groups, the Kruskal-Wallis test (nonparametric data) or one-way ANOVA (parametric data) test was applied to test for statistical significance. A *p*-value of less than 0.05 was considered to be statistically significant. Parametric data were presented as the mean and standard deviation (SD), whereas non-parametric data were presented as the median and interquartile range (IQR).

#### Mass spectrometer data analysis

The raw files generated by the mass spectrometer were imported into Proteome Discoverer v1.4 (Thermo Scientific) and processed using the Sequest and Amanda algorithms. Database interrogation was performed against the 2019-nCOVpFASTA1 database. Semi-tryptic cleavage with 2 missed cleavages was allowed for. Precursor mass tolerance was set to 10ppm and fragment mass tolerance set to 0.02 Da. Demamidation (NQ), oxidation (M) allowed as dynamic modifications. Peptide validation was performed using the Target-Decoy PSM validator node. The search results were imported into Scaffold Q+ for further validation (www.proteomesoftware.com) and statistical testing. A t-test was performed on the datasets and the emPAI quantitative method used to compare the datasets.

### Supplementary material and raw data

All supplementary material and raw data can be accessed here: https://1drv.ms/u/s!AgoCOmY3bkKHisg5J0nb6wqsBzzWAQ?e=XAsc7w

## RESULTS

### Fluorescence microscopy of purified fibrinogen and platelet poor plasma

Fluorescence microscopy was utilized to visualize the fluorescent amyloid signals in spontaneously formed anomalous clots, present in a fluorescent fibrinogen model, and also in healthy PPP with and without spike protein. As a preliminary investigation PPP from a single health control were incubated for 30 minutes with varying spike protein concentrations, followed by 30 minutes incubation with ThT and lastly preparation for viewing. It was found that the final exposure concentrations of 1 ng.mL^-1^ was sufficient and used for the rest of the study (see supplementary raw data for other exposure concentrations micrographs).

Figure 3A and B show representative micrographs of purified fluorescent (Alexa Fluor™488) fibrinogen with added thrombin and after exposure to 1 ng.mL^-1^ spike protein. A denser fibrin clot formed in the presence of spike protein (Figure 3B). In PPP, with and without thrombin, the green fluorescent ThT signal indicates areas of amyloid deposit formation. ThT is known to bind to open hydrophobic areas in protein and these are amyloid in nature (Adams et al., 2019, de Waal et al., 2018, Kell and Pretorius, 2017, Page et al., 2019). Figure 4A and D shows representative micrographs of a healthy PPP smear, with and without thrombin, and with added ThT where slight anomalous clotting is seen. In contrast, when spike protein is added to PPP, with and without thrombin, a major increase in dense anomalous clotted deposits, with an amyloid nature, were noted (referred to as amyloid deposits) (Figure 4B and D). A thresholding algorithm was applied to the micrographs (with and without thrombin) using Fiji^®^ which was used to calculate the total area of amyloid deposits in each micrograph (in total, 320 micrographs were analysed). Using this method, the average total percentage of amyloid deposits per group was calculated (naïve healthy PPP, naïve healthy PPP + thrombin, and PPP incubated with 1 ng.mL^-1^ spike protein, with and without added thrombin) (See Table 1). As expected, there were no significant differences between % area amyloid deposits of healthy PPP with and without thrombin However, there was a significant increase in % area amyloid deposits in PPP before and after added spike protein, in both PPP smears and fibrin clots (where thrombin was added).

**Table 1.** Percentage average amyloid area in platelet poor plasma (PPP) with and without spike protein and with and without thrombin.

| **Naïve healthy PPP vs naïve healthy PPP + added thrombin (n=10)** |  |
| --- | --- |
| P value (Wilcoxon test, paired non-parametric data expressed as median [Q1 – Q3]) | 0.2 |
| Median of naïve healthy samples | 0.3% [0.1% - 0.8%] |
| Median of naïve control samples (+ thrombin) | 0.9% [0.3% - 1.5%] |
| **Naïve healthy PPP + spike protein (1ng.mL<sup>-1</sup>) vs healthy PPP + spike protein (1ng.mL<sup>-1</sup>) + added thrombin) (n=10)** |  |
| P value (Data normally distributed; paired t-test) | 0.3 |
| Mean percentage amyloid of healthy samples + spike | 1.9% (± 1.2%) |
| Mean percentage amyloid of healthy samples + spike (+ Thrombin) | 2.4% (±1.3%) |
| **Naïve healthy PPP vs healthy PPP + spike protein (1ng.mL<sup>-1</sup>) (n=10)** |  |
| P value (Wilcoxon test, paired non-parametric data expressed as median [Q1 – Q3]) | **0.004 (\*\*)** |
| Median of healthy samples | 0.3% [0.1% - 0.8%] |
| Median of healthy samples + spike | 1.9% [1.2% - 2.4%] |
| **Naïve healthy PPP + added thrombin vs healthy PPP + spike protein (1ng.mL<sup>-1</sup>) + added thrombin) (n=10)** |  |
| P value (Data normally distributed; paired t-test) | **0.0036 (\*\*)** |
| Mean percentage amyloid of control samples (+ Thrombin) | 0.9% (± 0.6%) |
| Mean percentage amyloid of control samples + spike (+ Thrombin) | 2.4 % (1.3%) |

*Statistical significance was established at $p < 0.05$ . (\*\* = $p < 0,01$ ). Data is represented as either mean ± standard deviation or median [Q1 – Q3].* protein and with and without thrombin.

**Figure 3:**
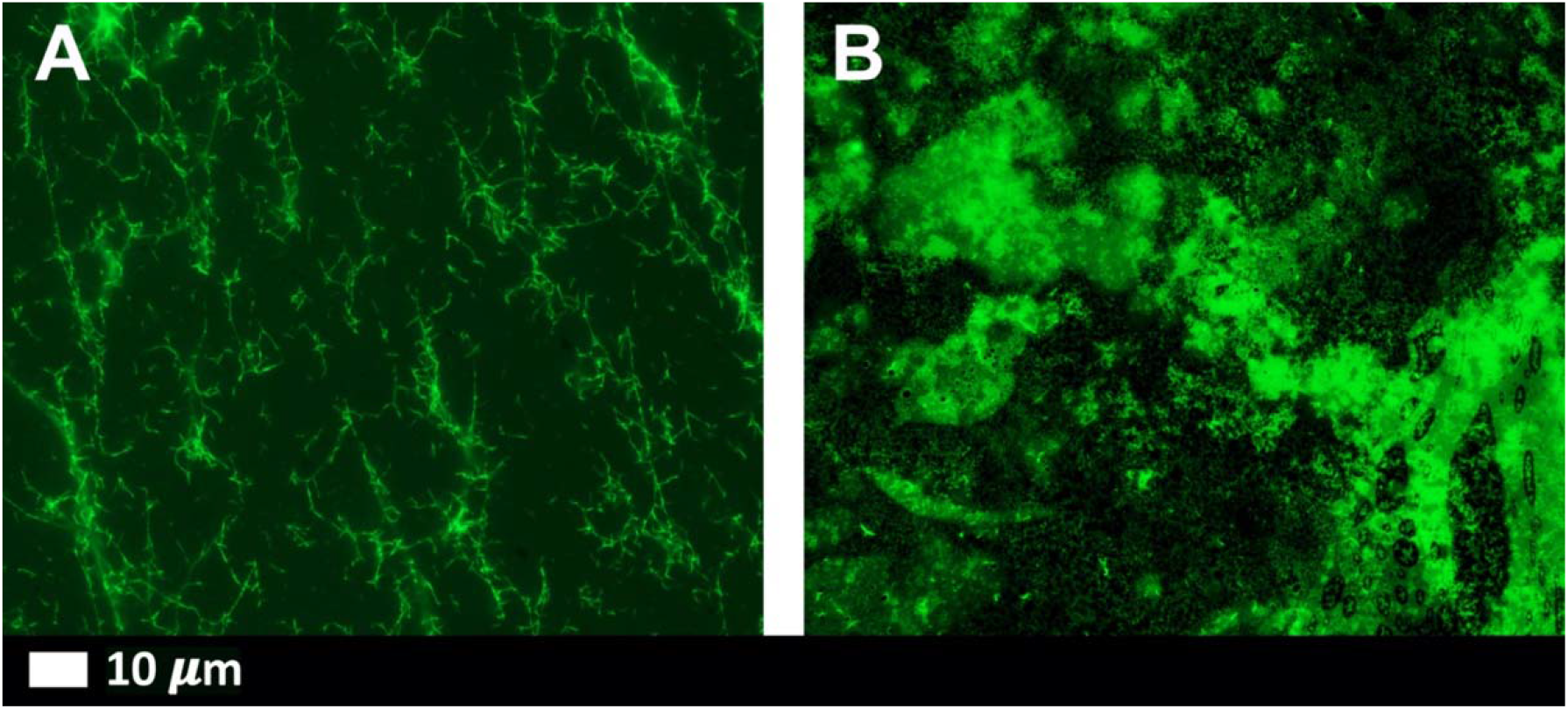
Representative fluorescence micrographs of purified fluorescent (Alexa Fluor™488) fibrinogen *(note no ThT added)* with added thrombin to form extensive fibrin clots. **A)** Fluorescent fibrinogen with thrombin; **B)** fluorescent fibrinogen with added spike protein (final exposure concentration 1 ng.mL^-1^) and thrombin.

**Figure 4:**
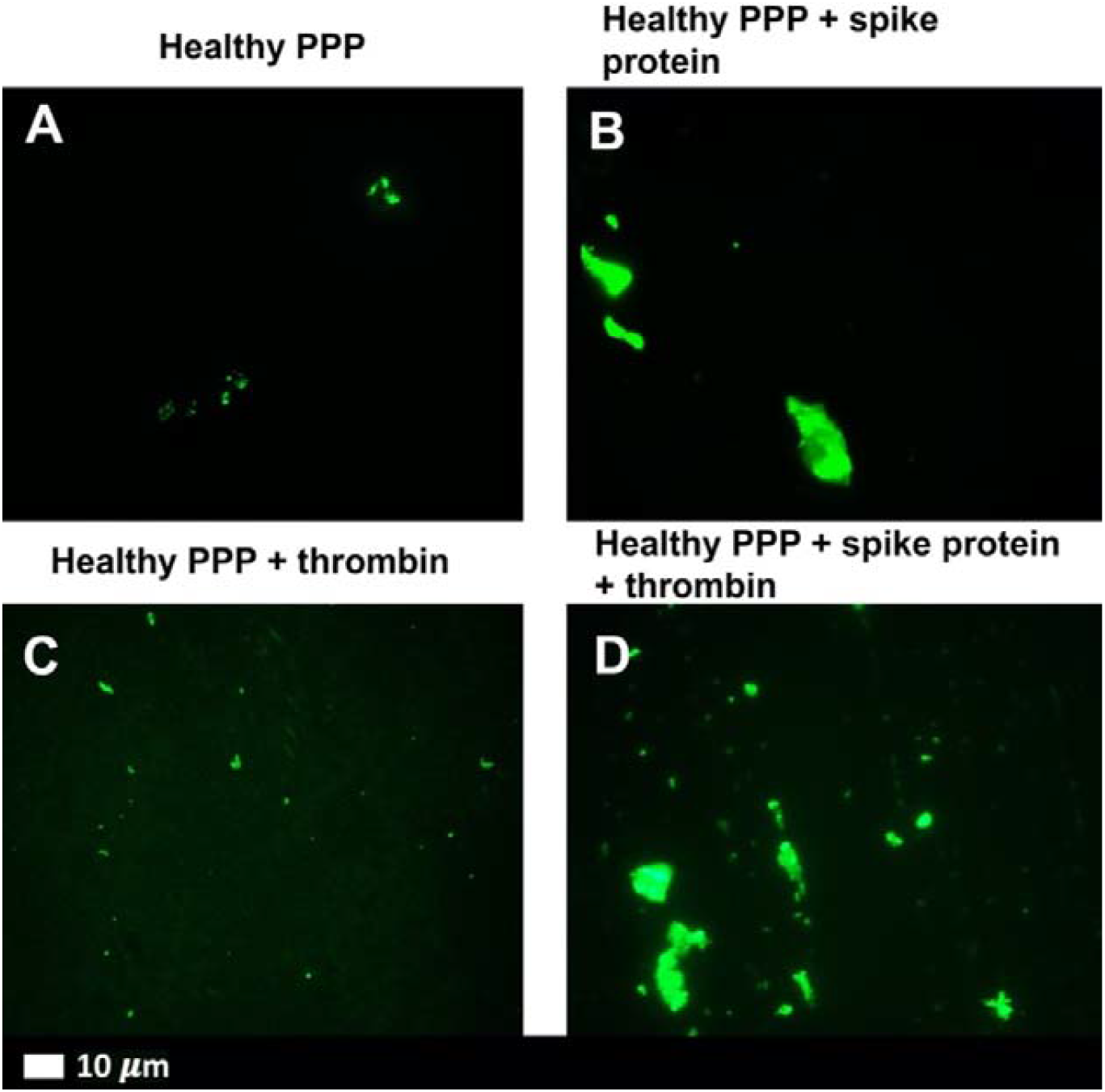
Representative fluorescence micrographs of platelet poor plasma (PPP) from healthy individuals after addition of ThT (green fluorescent signal). **A)** PPP smear. **B)** PPP with spike protein. **C)** PPP with thrombin to create extensive fibrin clot; **D)** PPP exposed to spike protein followed by addition of thrombin. Final spike protein concentration was 1ng.mL^-1^.

### Platelet activity

Fluorescence microscopy was used to visualize platelet activation in naive healthy WB and WB incubated with spike protein (1 ng.mL^-1^ final concentration). Samples were incubated with the platelet marker, CD62P-PE. Figure 5A shows representative platelets from naïve control samples, while Figure 5B show micrographs after spike protein incubation. Spike protein caused an increase in platelet hyperactivation (Figure 5B arrows).

**Figure 5A:**
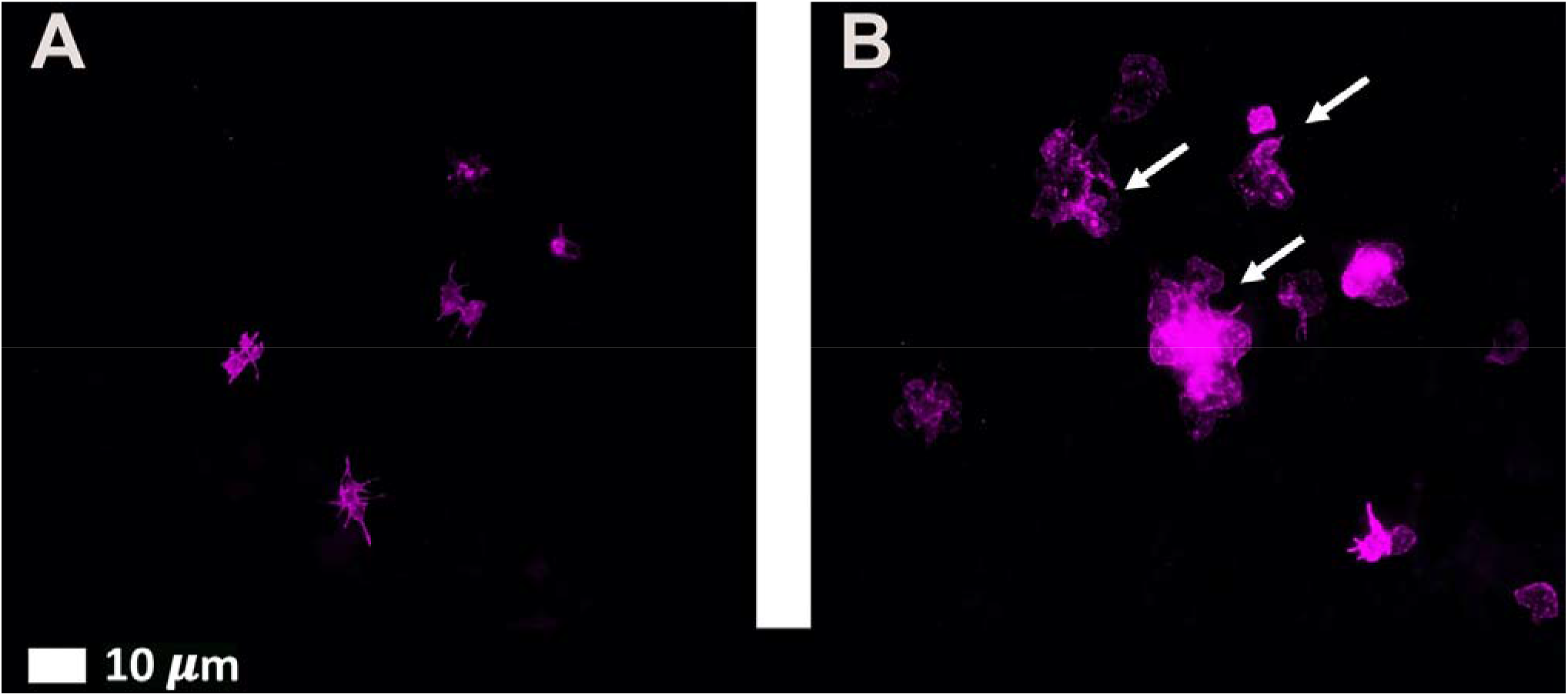
Fluorescence microscopy micrographs of representative naïve whole blood (WB), where platelets were incubated with fluorescent marker, CD62P-PE. **B)** WB after exposure to spike protein. The white arrows point to hyperactivated activated platelets.

### Scanning electron microscopy of whole blood

SEM was used to assess the erythrocyte and platelet ultrastructure after treatment with spike protein (1ng.mL^-1^ final concentration). Figure 6A and B shows micrographs from healthy WB samples, while Figure 6C to H shows micrographs of WB after incubation with spike protein. The majority of erythrocytes from healthy untreated controls were normocytic (regularly shaped) [Figure 6A (arrow)], and featured characteristic discoid shapes smooth, regular membrane surfaces. Slight platelet activation is seen due to contact activation (Figure 6B). The WB incubated with spike protein showed erythrocyte agglutination, despite the very low concentration of the spike protein. An increase in platelet hyperactivation, membrane spreading (Figure 6C and D), platelet-derived micro-particle formation were noted due to spike protein exposure. The formation of spontaneous and anomalous fibrin(ogen) deposits with an amyloid nature, were prominent in all the samples incubated with spike protein, without the addition of thrombin [Figure 6E to H (arrows)].

**Figure 6A to H:**
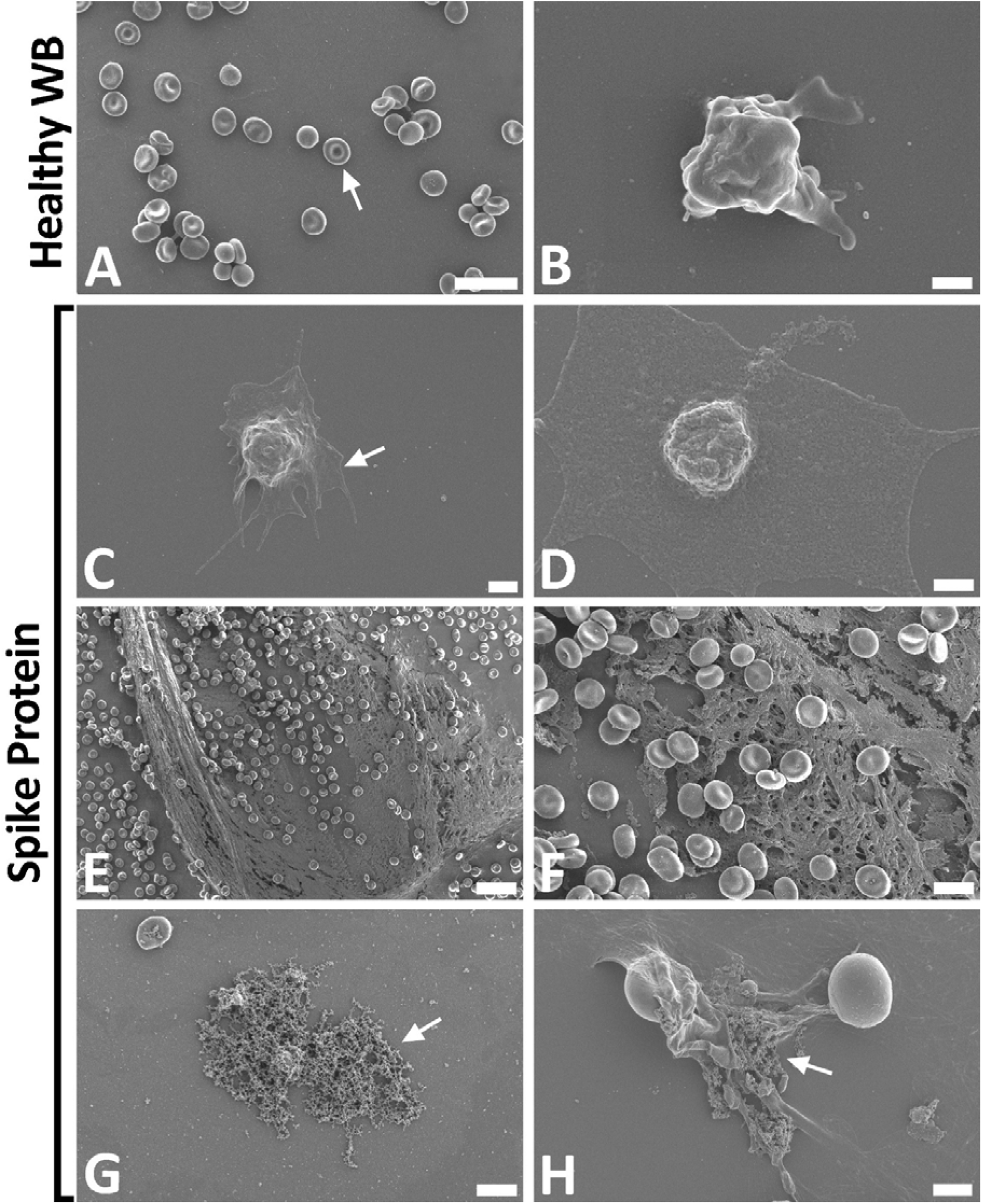
Representative scanning electron micrographs of healthy control whole blood (WB), with and without spike protein. **A and B)** Healthy WB smears, with arrow indicating normal erythrocyte ultrastructure. **C to H)** Healthy WB exposed to spike protein (1 ng.mL^-1^ final concentration), with **C and D**) indicating the activated platelets (arrow), **E and F**) showing the spontaneously formed fibrin network and **G and H**) the anomalous deposits that is amyloid in nature (arrows) (Scale bars: E: 20µm; A: 10µm; F and G: 5µm; H: 2µm; C: 1µm; B and D: 500nm).

### Microfluidics

Figure 7 show the clots that formed in the flow chambers after five minutes of starting the experiment. Healthy PPP formed a small clot along the bottom surface of the channel, as seen in Figure 7A. In healthy plasma, clot formation was a relatively slow and gradual process, resulting in the formation of a modest clot (see supplementary healthy PPP video 1). Clots formed in healthy PPP were relatively small and were limited to the walls of the flow channel. The clot had orderly layers that did not disrupt flow through the centre of the channel. As expected, clot formation was also less frequent than with the other samples (Figure 7B). The PPP with added spike protein showed a combination of a fibrous laminar clot and disorderly clotted mass (Figure 7E and F) (see supplementary healthy PPP with added spike protein video 2). The COVID-19 PPP show disorderly clots that cover the bulk of the channel and often protruding into the centre of the flow channel and disrupting flow (Figure 7C and D). In COVID-19 PPP, the reaction between thrombin and PPP occurred rapidly, resulting in large clots after approximately 90 seconds (see supplementary pooled COVID-19 patient PPP video 3). Interestingly, these clots did not propagate much after the initial burst, indicating that most of the thrombin was consumed in a short period of time. Clots also formed with the PPP with the addition of the spike protein, but not as disruptive as the COVID-19 PPP clots.

**Figure 7:**
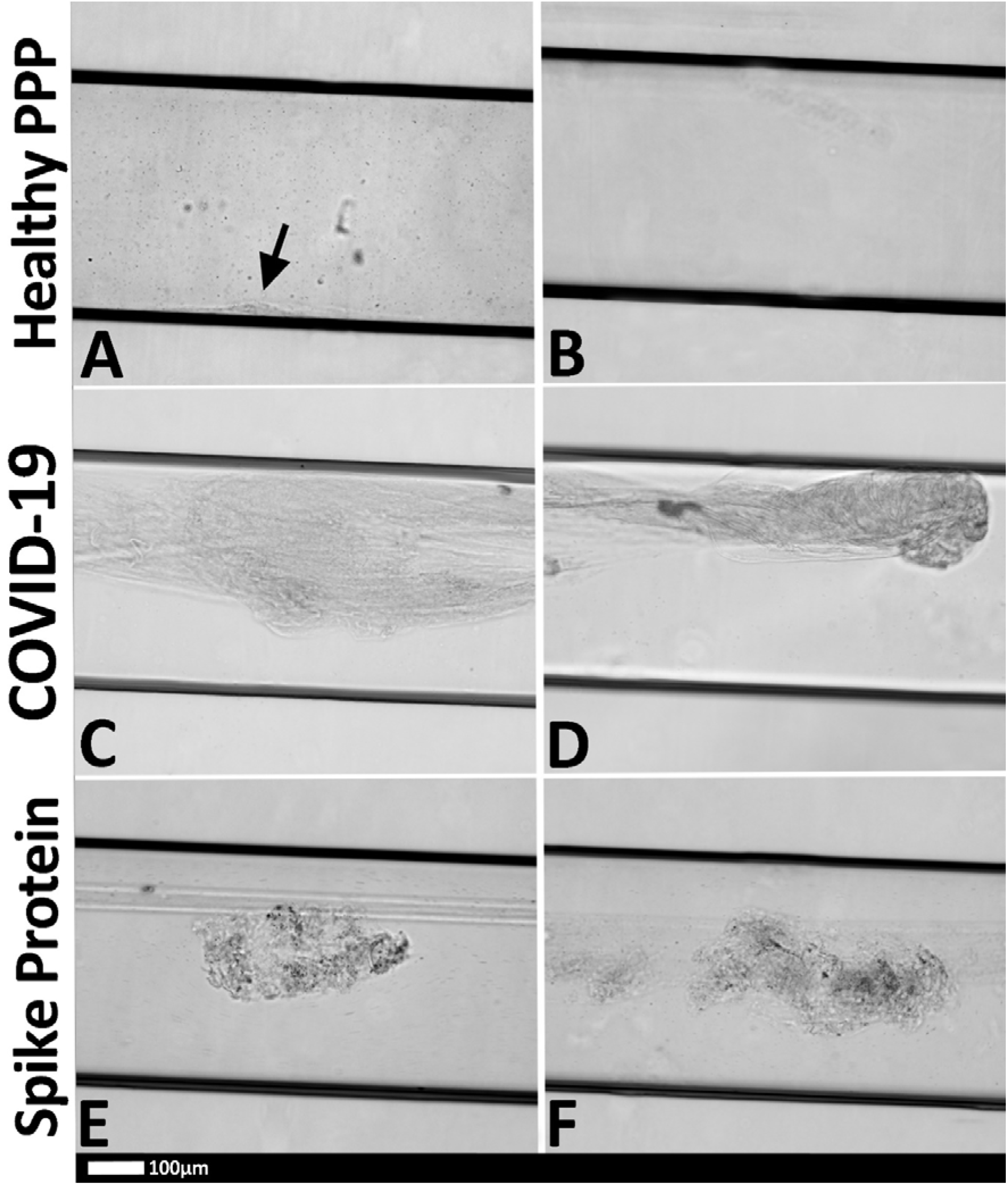
Representative micrographs of PPP clots in the microfluidic chambers (black horizontal lines are the outlines of the chambers) that were coated with thrombin. **A)** Healthy PPP clot, with small clot formation (arrow), with **B)** no clot formed in the healthy PPP sample; **C and D)** examples of clots from COVID-19 PPP samples and **E and F)** healthy PPP clot with spike protein.

An interesting observation was that clots from healthy PPP could easily be dislodged by flushing the flow channel with water at a rate of 1mL.min^-1^ (0.42m.s^-1^). Similarly, clots from healthy PPP with added spike protein could be dislodged in a similar fashion. COVID-19 clots, on the contrary, could not be displaced or dislodged and remained intact, even with the force of high-speed water flow in a small flow channel. This observation was consistent for all the COVID-19 samples.

### Mass spectrometry analysis

Figure 8 shows results from the mass spectrometry analysis. Mass spectrometry showed that when spike protein is added to healthy PPP, it results in structural changes to β and γ fibrin(ogen), complement 3 and prothrombin. These proteins were substantially resistant to trypsinization, in the presence of spike protein. (for sequence data see supplementary files).

**Figure 8:**
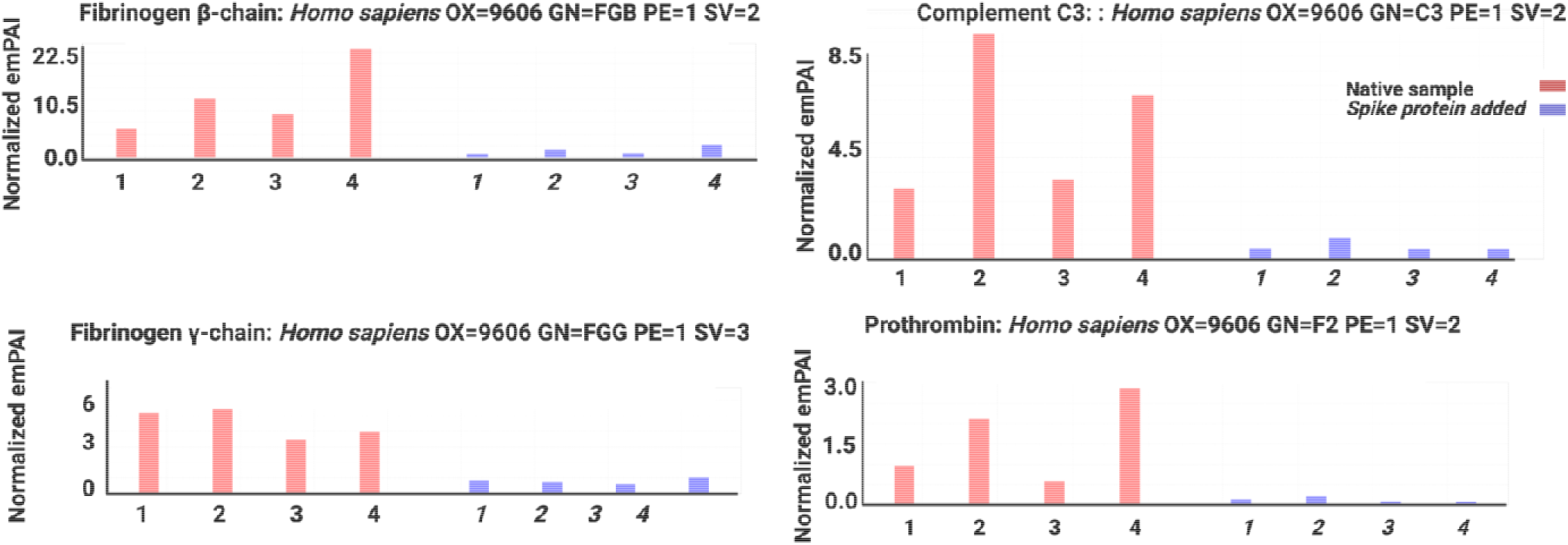
Mass spectrometry showing platelet poor plasma of 4 samples with and without added spike protein, Spike protein results in structural changes to β and γ fibrin(ogen), complement 3 and prothrombin.

## DISCUSSION

In this laboratory analysis, we provide evidence that spike protein does indeed play a major role in hypercoagulability seen in COVID-19 patients. It causes anomalous clotting in both purified fluorescent fibrinogen and in PPP, where the nature of the clots were shown to be amyloid (ThT as our amyloid dye of choice). An interesting observation was that these dense deposits were noted both in smears exposed to spike protein, and when thrombin was added. The addition of thrombin causes purified (Alexa Fluor™488) fibrinogen to polymerize into fibrin networks. Typically, these networks are netlike (Figure 3A). In the presence of spike protein, the structure changed to form dense clot deposits (Figure 3B). These deposits were seen in our fluorescent fibrin(ogen) model and PPP from healthy individuals exposed to spike protein. In healthy PPP exposed to spike protein, followed by incubation with ThT, there was a significant increase in anomalous clots with an amyloid nature, (Figure 4D), when compared to the health PPP. Spike protein also caused major ultrastructural changes in WB (as viewed with the SEM), where platelet hyperactivation were noted (Figure 6C and D). Increased in spontaneously formed fibrin network, as well as anomalous clot formation were also observed in SEM micrographs (Figure 6E-H). Interestingly, extensive spontaneous fibrin network formation was noted, <u>without</u> the addition of thrombin. This is in line with results that were recently published, where we showed similar ultrastructure in blood smears form COVID-19 positive patients. In these patient’s platelet hyperactivation, anomalous clotting with amyloid signal and spontaneous fibrin fibre formation were also observed (Pretorius et al., 2020, Venter et al., 2020).

With the microfluidic flow system, clots were formed, by infusing the entire microchannel with thrombin, thus simulating a hypercoagulable state, where endothelial damage was extensive. Given that the flow channel was made entirely of plastic and was devoid of any endothelial cells, the main component under investigation was the PPP (mostly fibrinogen protein) itself, which, in the case of the COVID-19 samples, may have contained downstream effects of some endothelial changes that would give rise to the hypercoagulable state that is characteristic of the disease. The flow setup used in this study could not directly account for endothelial changes but nonetheless demonstrated that COVID-19 also results in changes in the clotting profile of the PPP. This was evident in the rapid rate of thrombin consumption and fibrin formation in COVID-19 clots, and also in the nature of the PPP clots that were formed.

The clots that were observed in the healthy PPP with added spike protein, were of particular interest as they demonstrated a bridge between healthy PPP clots and COVID-19 clots. As described in the results, the healthy PPP clots were relatively small and orderly, while COVID-19 PPP clots were large, disorderly masses that formed rapidly and disrupted PPP flow in the channel. The healthy PPP clots with added spike protein, were a combination of the two, demonstrating disorderly clumped clot areas, co-existing with laminar fibrous PPP clots (which were larger than the healthy PPP clots). This intermediate state may arise from a number of factors, including the interaction of other biological actors which were absent from the flow setup and the time of exposure to spike protein. Further investigations would be beneficial for understanding the clotting mechanisms that are altered in the presence of spike protein.

One of the obvious differences, which was inadvertently observed while trying to clean the channels with high-speed water flow (i.e. by mechanical means), was the ease of healthy PPP and healthy PPP with added spike protein clot dissolution. However, there was a complete failure to dislodge or disturb COVID-19 PPP clots from the channels. Given the clot lysis and dissolution is a complex interplay between biochemical and biophysical factors, investigation of the effects of different therapeutic agents could elucidate this phenomenon (Hudson, 2017). The flow protocol used in this study would be a useful platform for testing different treatments for clinical application. A further limitation of this exploration is the use of PPP in investigating clot formation at a scale appropriate to the microvasculature. While the protocol enables the study of fibrin microclots, which are of interest in COVID-19, it excludes the influence of RBCs, which are known to heavily influence the non-Newtonian flow behaviour of blood at that scale (McHedlishvili, 1998). The inaccuracy of the flow regime arising from this exclusion and from the variability of viscosity introduces error into the results. Nonetheless, the inclusion of flow an appropriate spatial scale has enabled us to observe COVID-19 PPP clot formation over space and time, under dynamic conditions, and has given insights which would otherwise prove difficult to glean.

Mass spectrometry confirmed that spike protein causes structural changes to β and γ fibrin(ogen), complement 3 and prothrombin. These proteins become less resistant to trypsinization and changes the conformation, in such a way that there is a significant difference in peptide structure before and after spike protein addition.

## CONCLUSION

Scanning electron-and fluorescence microscopy revealed large dense anomalous and amyloid masses in whole blood and PPP of healthy individuals where spike protein was added to the samples. Mass spectrometry confirmed that when spike protein was added to PPP, it interacts with plasma proteins, resulting in fibrin(ogen), prothrombin and other proteins linked to coagulation, to become substantially resistant to trypsinization, resulting in less fragments. Flow analysis confirmed that microclots may impair blood flow. Here we suggest that, in part, the presence of spike protein in circulation may contribute to the hypercoagulation in COVID-19 positive patients and may cause severe impairment of fibrinolysis. Such lytic impairment may be the direct cause of the large microclots we have noted here in SEM and fluorescence microscopy, and previously in plasma samples of COVID-19 patients (Pretorius et al., 2020, Venter et al., 2020).

## Data Availability

https://1drv.ms/u/s!AgoCOmY3bkKHisg5J0nb6wqsBzzWAQ?e=XAsc7w

## DECLARATIONS

### Consent for publication

All authors approved submission of the paper.

## References

Ackermann, M., Verleden, S. E., Kuehnel, M., Haverich, A., Welte, T., Laenger, F., Vanstapel, A., Werlein, C., Stark, H., Tzankov, A., Li, W. W., Li, V. W., Mentzer, S. J. & Jonigk, D. 2020. Pulmonary Vascular Endothelialitis, Thrombosis, and Angiogenesis in Covid-19. N Engl J Med, 383, 120–128.

Adams, B., Nunes, J. M., Page, M. J., Roberts, T., Carr, J., Nell, T. A., Kell, D. B. & Pretorius, E. 2019. Parkinson’s Disease: A Systemic Inflammatory Disease Accompanied by Bacterial Inflammagens. Front Aging Neurosci, 11, 210.

Akhter, N., Ahmad, S., Alzahrani, F. A., Dar, S. A., Wahid, M., Haque, S., Bhatia, K., Sr Almalki, S., Alharbi, R. A. & Sindi, A. A. A. 2020. Impact of COVID-19 on the cerebrovascular system and the prevention of RBC lysis. Eur Rev Med Pharmacol Sci, 24, 10267–10278.

Bergmann, C. C. & Silverman, R. H. 2020. COVID-19: Coronavirus replication, pathogenesis, and therapeutic strategies. Cleve Clin J Med, 87, 321–327.

Berzuini, A., Bianco, C., Paccapelo, C., Bertolini, F., Gregato, G., Cattaneo, A., Erba, E., Bandera, A., Gori, A., Lamorte, G., Manunta, M., Porretti, L., Revelli, N., Truglio, F., Grasselli, G., Zanella, A., Villa, S., Valenti, L. & Prati, D. 2020. Red cell-bound antibodies and transfusion requirements in hospitalized patients with COVID-19. Blood, 136, 766–768.

Bobrova, L., Kozlovskaya, N., Korotchaeva, Y., Bobkova, I., Kamyshova, E. & Moiseev, S. 2020. Microvascular COVID-19 lung vessels obstructive thromboinflammatory syndrome (MicroCLOTS): a new variant of thrombotic microangiopathy? Crit Care Resusc, 22, 284.

Buzhdygan, T. P., Deore, B. J., Baldwin-Leclair, A., Mcgary, H., Razmpour, R., Galie, P. A., Potula, R., Andrews, A. M. & Ramirez, S. H. 2020. The SARS-CoV-2 spike protein alters barrier function in 2D static and 3D microfluidic in vitro models of the human blood–brain barrier. bioRxiv, 2020.06.15.150912.

Ciceri, F., Beretta, L., Scandroglio, A. M., Colombo, S., Landoni, G., Ruggeri, A., Peccatori, J., D’Angelo, A., De Cobelli, F., Rovere-Querini, P., Tresoldi, M., Dagna, L. & Zangrillo, A. 2020. Microvascular COVID-19 lung vessels obstructive thromboinflammatory syndrome (MicroCLOTS): an atypical acute respiratory distress syndrome working hypothesis. Crit Care Resusc, 22, 95–97.

De Waal, G. M., Engelbrecht, L., Davis, T., De Villiers, W. J. S., Kell, D. B. & Pretorius, E. 2018. Correlative Light-Electron Microscopy detects lipopolysaccharide and its association with fibrin fibres in Parkinson’s Disease, Alzheimer’s Disease and Type 2 Diabetes Mellitus. Sci Rep, 8, 16798.

Duan, L., Zheng, Q., Zhang, H., Niu, Y., Lou, Y. & Wang, H. 2020. The SARS-CoV-2 Spike Glycoprotein Biosynthesis, Structure, Function, and Antigenicity: Implications for the Design of Spike-Based Vaccine Immunogens. Front Immunol, 11, 576622.

Flores-Alanis, A., Sandner-Miranda, L., Delgado, G., Cravioto, A. & Morales-Espinosa, R. 2020. The receptor binding domain of SARS-CoV-2 spike protein is the result of an ancestral recombination between the bat-CoV RaTG13 and the pangolin-CoV MP789. BMC Res Notes, 13, 398.

George, S., Pal, A. C., Gagnon, J., Timalsina, S., Singh, P., Vydyam, P., Munshi, M., Chiu, J. E., Renard, I., Harden, C. A., Ott, I. M., Watkins, A. E., Vogels, C. B. F., Lu, P., Tokuyama, M., Venkataraman, A., Casanovas-Massana, A., Wyllie, A. L., Rao, V., Campbell, M., Farhadian, S. F., Grubaugh, N. D., Dela Cruz, C. S., Ko, A. I., Perez, A. B., Akaho, E. H., Moledina, D. G., Testani, J., John, A. R., Ledizet, M. & Mamoun, C. B. 2021. Evidence for SARS-CoV-2 Spike Protein in the Urine of COVID-19 patients. medRxiv, 2021.01.27.21250637.

Goshua, G., Pine, A. B., Meizlish, M. L., Chang, C. H., Zhang, H., Bahel, P., Baluha, A., Bar, N., Bona, R. D., Burns, A. J., Dela Cruz, C. S., Dumont, A., Halene, S., Hwa, J., Koff, J., Menninger, H., Neparidze, N., Price, C., Siner, J. M., Tormey, C., Rinder, H. M., Chun, H. J. & Lee, A. I. 2020. Endotheliopathy in COVID-19-associated coagulopathy: evidence from a single-centre, cross-sectional study. Lancet Haematol.

Grobler, C., Maphumulo, S. C., Grobbelaar, L. M., Bredenkamp, J. C., Laubscher, G. J., Lourens, P. J., Steenkamp, J., Kell, D. B. & Pretorius, E. 2020. Covid-19: The Rollercoaster of Fibrin(Ogen), D-Dimer, Von Willebrand Factor, P-Selectin and Their Interactions with Endothelial Cells, Platelets and Erythrocytes. Int J Mol Sci, 21.

Gupta, A., Madhavan, M. V., Sehgal, K., Nair, N., Mahajan, S., Sehrawat, T. S., Bikdeli, B., Ahluwalia, N., Ausiello, J. C., Wan, E. Y., Freedberg, D. E., Kirtane, A. J., Parikh, S. A., Maurer, M. S., Nordvig, A. S., Accili, D., Bathon, J. M., Mohan, S., Bauer, K. A., Leon, M. B., Krumholz, H. M., Uriel, N., Mehra, M. R., Elkind, M. S. V., Stone, G. W., Schwartz, A., Ho, D. D., Bilezikian, J. P. & Landry, D. W. 2020. Extrapulmonary manifestations of COVID-19. Nat Med, 26, 1017–1032.

Hudson, N. E. 2017. Biophysical Mechanisms Mediating Fibrin Fiber Lysis. Biomed Res Int, 2017, 2748340.

Jacob, M., Chappell, D. & Becker, B. F. 2016. Regulation of blood flow and volume exchange across the microcirculation. Crit Care, 20, 319.

Kawase, M., Kataoka, M., Shirato, K. & Matsuyama, S. 2019. Biochemical Analysis of Coronavirus Spike Glycoprotein Conformational Intermediates during Membrane Fusion. J Virol, 93.

Kell, D. B. & Pretorius, E. 2017. Proteins behaving badly. Substoichiometric molecular control and amplification of the initiation and nature of amyloid fibril formation: lessons from and for blood clotting. Prog Biophys Mol Biol, 123, 16–41.

Lam, L. M., Murphy, S. J., Kuri-Cervantes, L., Weisman, A. R., Ittner, C. G., Reilly, J. P., Pampena, M. B., Betts, M. R., Wherry, E. J., Song, W. C., Lambris, J. D., Cines, D. B., Meyer, N. J. & Mangalmurti, N. S. 2020. Erythrocytes Reveal Complement Activation in Patients with COVID-19. medRxiv.

Letarov, A. V., Babenko, V. V. & Kulikov, E. E. 2020. Free SARS-CoV-2 Spike Protein S1 Particles May Play a Role in the Pathogenesis of COVID-19 Infection. Biochemistry (Moscow).

Mchedlishvili, G. 1998. Disturbed blood flow structuring as critical factor of hemorheological disorders in microcirculation. Clin Hemorheol Microcirc, 19, 315–25.

Page, M. J., Thomson, G. J. A., Nunes, J. M., Engelbrecht, A. M., Nell, T. A., De Villiers, W. J. S., De Beer, M. C., Engelbrecht, L., Kell, D. B. & Pretorius, E. 2019. Serum amyloid A binds to fibrin(ogen), promoting fibrin amyloid formation. Sci Rep, 9, 3102.

Perico, L., Benigni, A., Casiraghi, F., Ng, L. F. P., Renia, L. & Remuzzi, G. 2021. Immunity, endothelial injury and complement-induced coagulopathy in COVID-19. Nat Rev Nephrol, 17, 46–64.

Pretorius, E. 2013. The adaptability of red blood cells. Cardiovasc Diabetol, 12, 63.

Pretorius, E., Bester, J., Mbotwa, S., Robinson, C. & Kell, D. B. 2016. Acute induction of anomalous blood clotting by molecular amplification of highly substoichiometric levels of bacterial lipopolysaccharide (LPS). BioRxiv, 053538.

Pretorius, E., Oberholzer, H. M., Van Der Spuy, W. J. & Meiring, J. H. 2010. Smoking and coagulation: the sticky fibrin phenomenon. Ultrastruct Pathol, 34, 236–9.

Pretorius, E., Venter, C., Laubscher, G. J., Lourens, P. J., Steenkamp, J. & Kell, D. B. 2020. Prevalence of readily detected amyloid blood clots in ‘unclotted’ Type 2 Diabetes Mellitus and COVID-19 plasma: a preliminary report. Cardiovasc Diabetol, 19, 193.

Pretorius, E., Vermeulen, N., Bester, J. & Lipinski, B. 2013a. Novel use of scanning electron microscopy for detection of iron-induced morphological changes in human blood. Microsc Res Tech, 76, 268–71.

Pretorius, E., Vermeulen, N., Bester, J., Lipinski, B. & Kell, D. B. 2013b. A novel method for assessing the role of iron and its functional chelation in fibrin fibril formation: the use of scanning electron microscopy. Toxicol Mech Methods, 23, 352–9.

Renzi, S., Landoni, G., Zangrillo, A. & Ciceri, F. 2020. MicroCLOTS pathophysiology in COVID 19. Korean J Intern Med.

Rhea, E. M., Logsdon, A. F., Hansen, K. M., Williams, L. M., Reed, M. J., Baumann, K. K., Holden, S. J., Raber, J., Banks, W. A. & Erickson, M. A. 2021. The S1 protein of SARS-CoV-2 crosses the blood–brain barrier in mice. Nature Neuroscience, 24, 368–378.

Roberts, I., Muelas, M. W., Taylor, J. M., Davison, A. S., Xu, Y., Grixti, J. M., Gotts, N., Sorokin, A., Goodacre, R. & Kell, D. B. 2020. Untargeted metabolomics of COVID-19 patient serum reveals potential prognostic markers of both severity and outcome. medRxiv, 2020.12.09.20246389.

Venter, C., Bezuidenhout, J. A., Laubscher, G. J., Lourens, P. J., Steenkamp, J., Kell, D. B. & Pretorius, E. 2020. Erythrocyte, Platelet, Serum Ferritin, and P-Selectin Pathophysiology Implicated in Severe Hypercoagulation and Vascular Complications in COVID-19. Int J Mol Sci,

Walls, A. C., Park, Y. J., Tortorici, M. A., Wall, A., Mcguire, A. T. & Veesler, D. 2020. Structure, Function, and Antigenicity of the SARS-CoV-2 Spike Glycoprotein. Cell, 181, 281–292.e6.

Watanabe, Y., Allen, J. D., Wrapp, D., Mclellan, J. S. & Crispin, M. 2020. Site-specific glycan analysis of the SARS-CoV-2 spike. Science, 369, 330–333.

Zhang, C., Zheng, W., Huang, X., Bell, E. W., Zhou, X. & Zhang, Y. 2020. Protein structure and sequence re-analysis of 2019-nCoV genome does not indicate snakes as its intermediate host or the unique similarity between its spike protein insertions and HIV-1. bioRxiv.

